# Heart Rate Variability Based Ventilatory Threshold Estimation – Validation of a Commericially Available Algorithm

**DOI:** 10.1101/2024.08.14.24311967

**Authors:** Timo Eronen, Jukka A. Lipponen, Vesa V. Hyrylä, Saana Kupari, Jaakko Mursu, Mika Venojärvi, Heikki O. Tikkanen, Mika P. Tarvainen

**Affiliations:** School of Medicine, Institute of Biomedicine, University of Eastern Finland, Kuopio, Finland; Department of Technical Physics, University of Eastern Finland, Kuopio, Finland; Department of Emergency Care, Kuopio University Hospital; Department of Clinical Physiology and Nuclear Medicine, Kuopio University Hospital, Kuopio, Finland

**Keywords:** Detrended fluctuation analysis, Exercise testing, Heart rate variability, CPET, ventilatory threshold

## Abstract

Ventilatory thresholds (VT1 and VT2) are critical in exercise prescription and athletic training, delineating the transitions from aerobic to anaerobic metabolism. More specifically, VT1 signifies the onset of lactate accumulation whilst VT2 signifies the onset of metabolic acidosis. Accurate determination of these thresholds is vital for optimizing training intensity. Fractal correlation properties of heart rate variability (HRV), particularly the short-term scaling exponent alpha 1 of Detrended Fluctuation Analysis (DFA-α1), have demonstrated potential for this purpose. This study validates the accuracy of commercial ventilatory threshold estimation algorithm (VT-algorithm) developed by Kubios. The VT-algorithm employs instantaneous heart rate (HR) relative to HR reserve and respiratory rate (RF), along with the DFA-α1. Sixty-four physically active participants underwent an incremental cardiopulmonary exercise test (CPET) with inter-beat interval (RR) measurements. DFA-α1 and the Kubios VT-algorithm were used to assess HR and oxygen uptake (VO2) at ventilatory thresholds. On average VO2 at true VT, DFA-α1, and VT-algorithm derived ventilatory thresholds were 1.74, 2.00 and 1.89 l/min (VT1) and 2.40, 2.41 and 2.40 l/min (VT2), respectively. Correspondingly, average HRs at the true VT, DFA-α1, and VT-algorithm thresholds were 141, 151 and 142 bpm (VT1) and 169, 168 and 170 bpm (VT2), respectively. When compared to the true thresholds, Bland-Altman error statistics (bias ± standard deviation of error) for the DFA-α1 thresholds were -0.26±0.41 l/min or -10±16 bpm at VT1 and 0.00±0.34 l/min or 1±10 bpm at VT2, whereas the VT-algorithm errors were - 0.15±0.28 l/min or -1±11 bpm at VT1 and 0.01±0.20 l/min or -1±7 bpm at VT2. HRV based VT determination algorithms accurately estimate ventilatory thresholds, offering insights into training zones, internal loading, and metabolic transitions during exercise without the need of laboratory equipment. The Kubios VT-algorithm, which incorporates instantaneous HR and RF along with DFA-α1, provided higher accuracy for VO2 and HR values for both VT1 and VT2.

## INTRODUCTION

Ventilatory thresholds (VT1 and VT2) are often used in exercise prescription and athletic training to define training intensities and training zones. Increased lactate accumulation causes metabolic acidosis which is subdued by body’s buffering systems (e.g. bicarbonate-CO2 buffering system). At VT2 body’s buffering systems cannot keep up with increasing metabolic acidosis, leading to a more significant reliance on anaerobic metabolism. (1) Ventilatory thresholds can be determined by the following two methods. Lactate measurements can be used to identify turning points in lactate production as lactate thresholds. Another approach to determine ventilatory thresholds is through cardiopulmonary exercise testing (CPET). Ventilatory thresholds can be determined from respiratory gases via CPET by addressing increases in minute ventilation and its relationship to oxygen uptake (VO2) and carbon dioxide production (VCO2).

Heart rate variability (HRV) based ventilatory threshold detection methods have been under study for the past 25 years (2). As exercise intensity increases, the autonomic nervous system gradually shifts from parasympathetic to sympathetic dominance (3). Dimensionless HRV indexes based on fractal correlation properties, such as the short-term scaling exponent alpha 1 of Detrended Fluctuation Analysis (DFA-α1), have a broad dynamic range encompassing the low, moderate and high exercise intensity domains (4,5). DFA-α1 is derived from the fractal arrangement of heartbeat intervals over short time spans. Increasing sympathetic activity and parasympathetic withdrawal leads to changes in HRV, including DFA-α1 (6). At low intensity exercise (below VT1), DFA-α1 values typically range from 1.0 to 0.75, and thereby, VT1 is often identified when DFA-α1 reaches 0.75 (7). As exercise intensity continues to increase, DFA-α1 decreases further with a value of 0.5 representing VT2 (8). Beyond this, during the most intense exercise, DFA-α1 drops below 0.5. The shift in DFA-α1 from 1.0 towards 0.5 indicates a transition in the correlation properties of HRV data, moving from a fractal behavior typically seen during rest to behavior resembling random white noise (9).

The utilisation of DFA-α1 in assessing ventilatory thresholds has significant attention in the fields of exercise prescription and the evaluation of endurance exercise fatigue. However, DFA-α1 has limitations and potential pitfalls. The most influential factors affecting successful DFA-α1 measurement are device-related errors, such as low signal-to-noise ratio due to suboptimal electrode placement or low beat detection precision (10). Kubios HRV is a widely used software for evaluating HRV and accessing DFA-α1 (11). The software also includes a validated algorithm for correcting abnormal beat intervals, which improves the accuracy of HRV metrics including DFA-α1 (12). The accuracy of the DFA-α1-based ventilatory threshold estimates, compared to the thresholds determined from CPET, is often assessed through correlation of either heart rate (HR) or VO2 at the estimated and reference thresholds. Previous studies have reported varying correlations for HR, ranging from 0.66 to 0.87 for VT1 and 0.71 to 0.90 for VT2 (13–15) and for VO2 0.66 for VT1 and 0.74 for VT2 (13).

The aim of this study was to validate the accuracy of the ventilatory threshold estimation algorithm available in the Kubios HRV Scientific software (Kubios Oy, Kuopio, Finland). This algorithm utilizes heart rate and HRV based estimate of respiratory rate along with DFA-α1 in the estimation of ventilatory thresholds. HR reserve has been associated with oxygen uptake reserve at ventilatory thresholds as shown by Gaskill et al. (16), and relationship between respiratory frequency (RF) and ventilatory thresholds were proposed by Cross et al. (17). Furthermore, a recent study from Rogers et al. showed that using DFA-α1 together with RF improves the accuracy of VT estimates when compared to DFA-α1 alone (18). Thus, we hypothesized that the accuracy of ventilatory threshold determination could be significantly improved by including these additional physiological measures, which are both known to correlate with exercise intensity and metabolic demand, in the determination of ventilatory thresholds.

## MATERIALS & METHODS

### Participants

Sixty-four recreationally active voluntary men and women, whose subject demographics are presented in Table 1, participated in two different study protocols, both of which included identical exercise testing measurements. Prior to testing, participants were instructed to abstain from caffeine, alcohol, tobacco, and vigorous exercise for 24 hours and no infection symptoms for two weeks. The studies were conducted in compliance with the principles of the Declaration of Helsinki, and they were approved by the Ethics Committee of the Northern Savo Hospital District in Kuopio, Finland (484/13.00.00/2017 and 409/13.02.00/2019). Participants provided their written informed consent before participating in the study.

**Table 1a:** MALE Participant characteristics.

| N = 32 | MEAN ± SD | Min | Max |
| --- | --- | --- | --- |
| Age (years) | 34.7 ± 6.4 | 23 | 48 |
| Height (cm) | 179.5 ± 5.8 | 170 | 194 |
| Weight (kg) | 80.3 ± 9.4 | 60.0 | 96.8 |
| VO2 max (l/min) | 3.41 ± 0.60 | 2.28 | 4.41 |
| HR max (bpm) | 186 ± 10 | 173 | 212 |

**Table 1b:** FEMALE Participant characteristics.

| N = 32 | MEAN ± SD | Min | Max |
| --- | --- | --- | --- |
| Age (years) | 32.5 ± 8.7 | 20 | 50 |
| Height (cm) | 167.8 ± 6.1 | 157 | 180 |
| Weight (kg) | 69.1 ± 12.0 | 54.2 | 98.7 |
| VO2 max (l/min) | 2.47 ± 0.39 | 1.69 | 3.32 |
| HR max (bpm) | 185 ± 11 | 167 | 204 |

Abbreviations: VO2max, maximal oxygen uptake; HRmax, maximum heart rate.

### Exercise testing

The subjects underwent an incremental cardiopulmonary exercise test (CPET) on an ergometer (Ergoselect 100, Ergoline GmbH, Germany) in a temperature-controlled room. The exercise protocol began with a seated 5-minute rest period on the ergometer, after which the incremental protocol was initiated. For men, this protocol started at 35 W and increased 35 W every 3 minutes, while for women the start was 25 W and increase was set at 25 W. The test was performed to volitional maximal effort and was terminated if the participant could not maintain a cadence of over 60 rpm. Verbal encouragement was provided to help participants achieve maximal effort. Maximal effort was achieved if the participant reached over 95% of predicted HRmax, an over 1.1 respiratory exchange ratio (RER) was observed at peak exercise, or a plateau in oxygen uptake was observed. Heart rate and ECG channel were continuously measured during the exercise test using the Bittium Faros or ME6000 biomonitor (Bittium Ltd, Finland). Respiratory gases and ventilatory measurements were collected using the breath-by-breath method with the Cortex Metamax 3b portable spiroergometry device (Cortex Biophysik GmbH, Germany). Exercise protocol, along with HR, RF, DFA-α1, and VT-algorithm signals for a representative study participant are shown in Figure 1.

**Figure 1:**
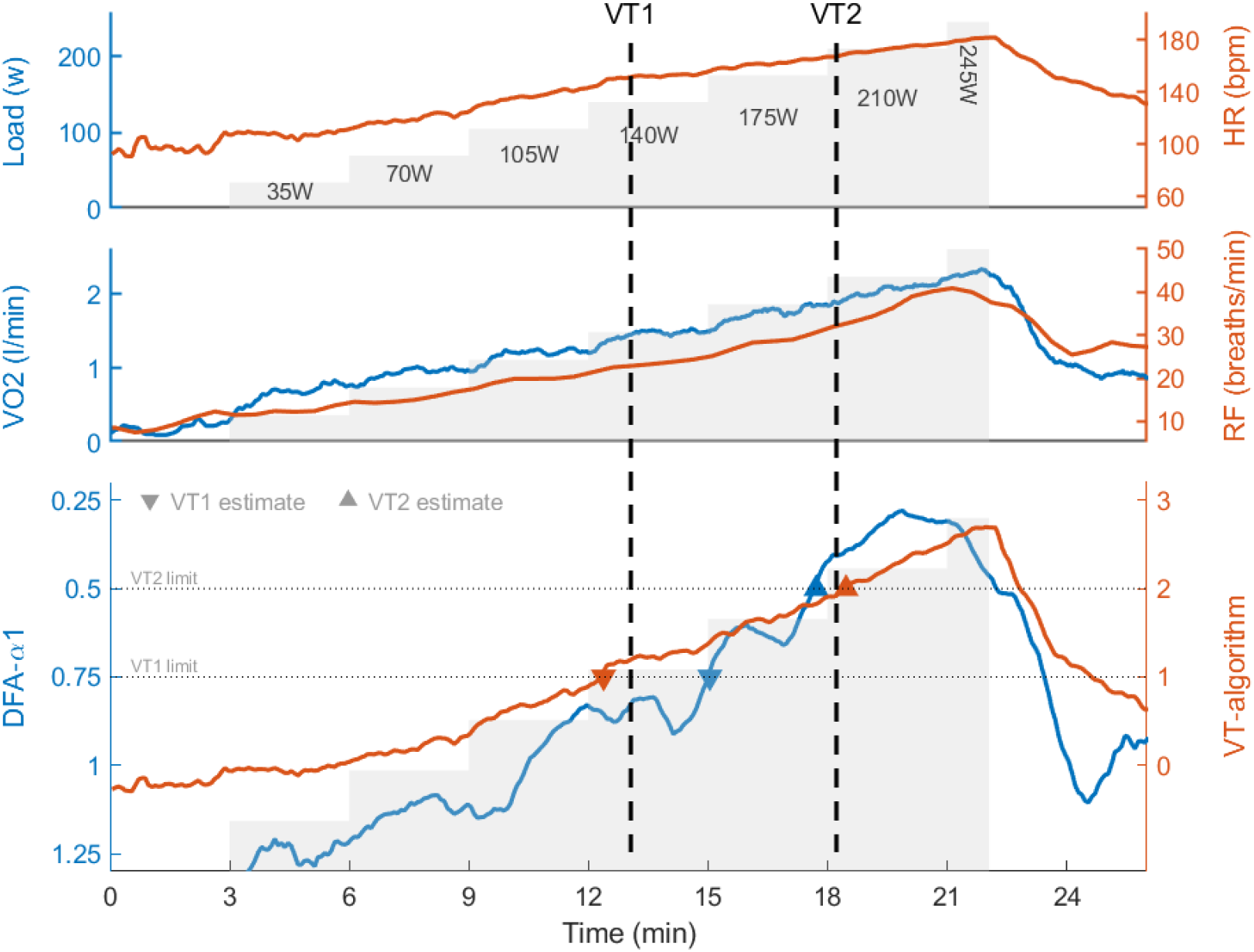
Illustration of the exercise protocol as well as heart rate, oxygen uptake, respiratory rate, DFA-α1, and VT-algorithm signals for a representative study participant. True ventilatory thresholds are marked by vertical dashed lines, and the VT estimates obtained from the DFA-α1 (red) and VT-algorithm (blue) are marked with triangle markers.

### VT Determination

Ventilatory thresholds were analysed from the CPET data (later referred as true VT) using combination of criteria chosen by an experienced exercise physiology specialist (TE). To confirm the ventilatory threshold an independent researcher was involved. For VT1 determination, a dual method approach was employed, involving an observed increase in VCO2/VO2 slope (the modified V-slope method) with a nadir or turning point in VE/VO2 equivalent curve, coinciding with an RER approaching 1.0, as described in (19). For VT2 determination, the criteria included an increase in the VE/VCO2 slope, a deflection of PETCO2, and an RER value of 1.0 (20). There was a good agreement of ventilatory thresholds determined between two researchers. Mean differences for VT1 and VT2 were -0.004 l/min (±0.01) and 0.007 l/min (±0.01), respectively. The interclass correlation coefficients (ICC) for VT1 was 0.990 (CI 0.983-0.994) and for VT2 0.995 (CI 0.992-0.997), showing excellent agreement between two researchers.

### DFA alpha 1

ECG data was processed using Kubios HRV Scientific software version 4.0. The software automatically applied beat detection, noise detection, and beat correction to the ECG data. Nevertheless, visual inspection of the entire test recordings was performed to ensure the exclusion of any poor-quality (less than 5% correction in beats) data segments or arrhythmia episodes from the analysis. The very low frequency trend components were removed from the RR interval data using the smoothness priors method, with a smoothing parameter set to 500 (21). For the computation of the DFA-α1 parameter, a range of 4-16 beats was utilized to capture the fractal correlation of RR interval short-term DFA-α1. DFA-α1 was calculated within a moving window, with the window width set to 120 seconds and the window shifted in 5-second intervals. Ventilatory thresholds based on DFA-α1 were determined by plotting DFA-α1 against time, identifying the time points at which DFA-α1 crossed the values of 0.75 (VT1) or 0.5 (VT2), and finally extracting the HR and VO2 values at these time points (see Figure 1). One participant was excluded from the DFA-α1 analysis due to consistently high (> 0.5) DFA-α1 value and therefore not reaching VT2 by solely using DFA-α1 analysis.

### VT-algorithm

In addition to the DFA-α1 approach, the ventilatory threshold estimation algorithm (VT-algorithm) available in the Kubios HRV Scientific software was employed to determine both ventilatory thresholds. This algorithm relies on the instantaneous values of heart rate in relation to HR reserve, HRV based respiratory rate estimate (RF), and the fractal behaviour of HRV measured by DFA-α1. The HR reserve (HRR) was obtained by subtracting a fixed 60 bpm resting HR from the maximal HR achieved during the CPET. The RF was derived from beat-to-beat RR data within a 30-second moving window as described in (22). Ventilatory threshold estimates were determined by plotting the VT-algorithm output against time, identifying the points at which the VT-algorithm output crossed the values of 1 (VT1) or 2 (VT2), and then taking the HR and VO2 values at these time points (see Figure 1).

### Statistical analysis

Descriptive and analytical statistics were performed using IBM SPSS Statistics 27. The agreement of threshold values was assessed via linear regression, Pearson’s r correlation coefficient, standard error of estimate (SEE) and Bland-Altman plots with limits of agreement (23). The Pearson’s r correlations were interpret as follows: 0.3 < r < 0.5 moderate, 0.5 ≤ r < 0.7 high, 0.7 ≤ r < 0.9 very high and r ≥ 0.9 almost perfect (24). Paired t-testing was used for the comparison of true VTs vs. DFA-α1 estimate and VT-algorithm. For all tests, the statistical significance was accepted as p < 0.05. The interpretation of effect sizes is based on Cohen’s thresholds for small effects (d < 0.5), moderate effects (d ≥ 0.5) and large effects (d > 0.8)(25). Additionally, ICC was calculated to assess the inter-researcher agreement in VT determination.

## RESULTS

The main characteristics of the study participants are shown in Table 1 (a and b). The statistical comparison between the true ventilatory thresholds and the ventilatory thresholds obtained from DFA-α1 and VT-algorithm is presented comprehensively in Table 2. Furthermore, the Bland-Altman plots illustrating the bias and error of the VT estimates regarding HR and VO2 are provided in Figures 2 and 3, respectively. Regarding HR at VT1, DFA-α1 exhibited a moderate correlation (r=0.44, standard error of estimation, SEE 13.18), but the mean HR value was significantly over-estimated when compared to the HR at the true VT1 (151 vs. 141 bpm, p<0.001, d=0.55). In contrast, the VT-algorithm demonstrated a high correlation (r=0.62, SEE 11.37), and the mean HR value was like the HR at the true VT1 (142 vs. 141 bpm, p=0.93, d=0.01). For HR at VT2, DFA-α1 showed a high correlation (r=0.66, SEE 9.40), and VT-algorithm had a very high correlation (r=0.82, SEE 6.20). The mean HRs at the estimated VT2 were like the HR at the true VT2 for both DFA-α1 (168 vs. 169 bpm, p=0.60, d=0.07) and VT-algorithm (170 vs. 169 bpm, p=0.58, d=0.07). The bias and standard deviation of the error for DFA-α1, obtained from the Bland-Altman analysis, were 10±18 bpm at VT1 and 1±10 bpm at VT2. For VT-algorithm, the errors were 1±11 bpm at VT1 and 1±7 bpm at VT2.

**Table 2:** Comparison between true ventilatory thresholds (True VT) and ventilatory thresholds obtained by Detrend Fluctuation Analysis (DFA-α1) and Ventilatory threshold detection algorithm (VT-algorithm).

| | True VT value<br>MEAN ± SD | DFA-α1 estimate<br>MEAN ± SD | VT-algorithm<br>MEAN ± SD |
| --- | --- | --- | --- |
| VT1 HR (bpm) | 144 ± 14 | 151 ± 19 | 142 ± 8 |
| VT1 HR error | | -10 ± 18* | -1 ± 11 |
| VT1 HR r |  | 0.44** | 0.62** |
| VT2 HR (bpm) | 169 ± 12 | 168 ± 13† | 170 ± 9 |
| VT2 HR error | | 1 ± 10 | -1 ± 7 |
| VT2 HR r |  | 0.66** | 0.82** |
| VT1 VO<sub>2</sub> (l/min) | 1.74 ± 0.41 | 2.00 ± 0.55 | 1.89 ± 0.48 |
| VT1 VO<sub>2</sub> error | | -0.26 ± 0.41* | -0.15 ± 0.28* |
| VT1 VO<sub>2</sub> r |  | 0.67** | 0.81** |
| VT2 VO<sub>2</sub> (l/min) | 2.40 ± 0.56 | 2.41 ± 0.57† | 2.40 ± 0.55 |
| VT2 VO<sub>2</sub> error | | -0.00 ± 0.34 | 0.01 ± 0.20 |
| VT2 VO<sub>2</sub> r |  | 0.82** | 0.93** |

† One subject excluded, DFA-α1 was higher than 0.5 during whole recording
\* Mean value statistically different to the value at the true threshold (t-test, $p<0.05$ )
\*\* Statistically significant correlation ( $p<0.001$ )
Abbreviations: VT1, first ventilatory threshold; VT2, second ventilatory threshold; HR, heart rate; VO<sub>2</sub>, oxygen consumption; r, Pearson correlation value

**Figure 2:**
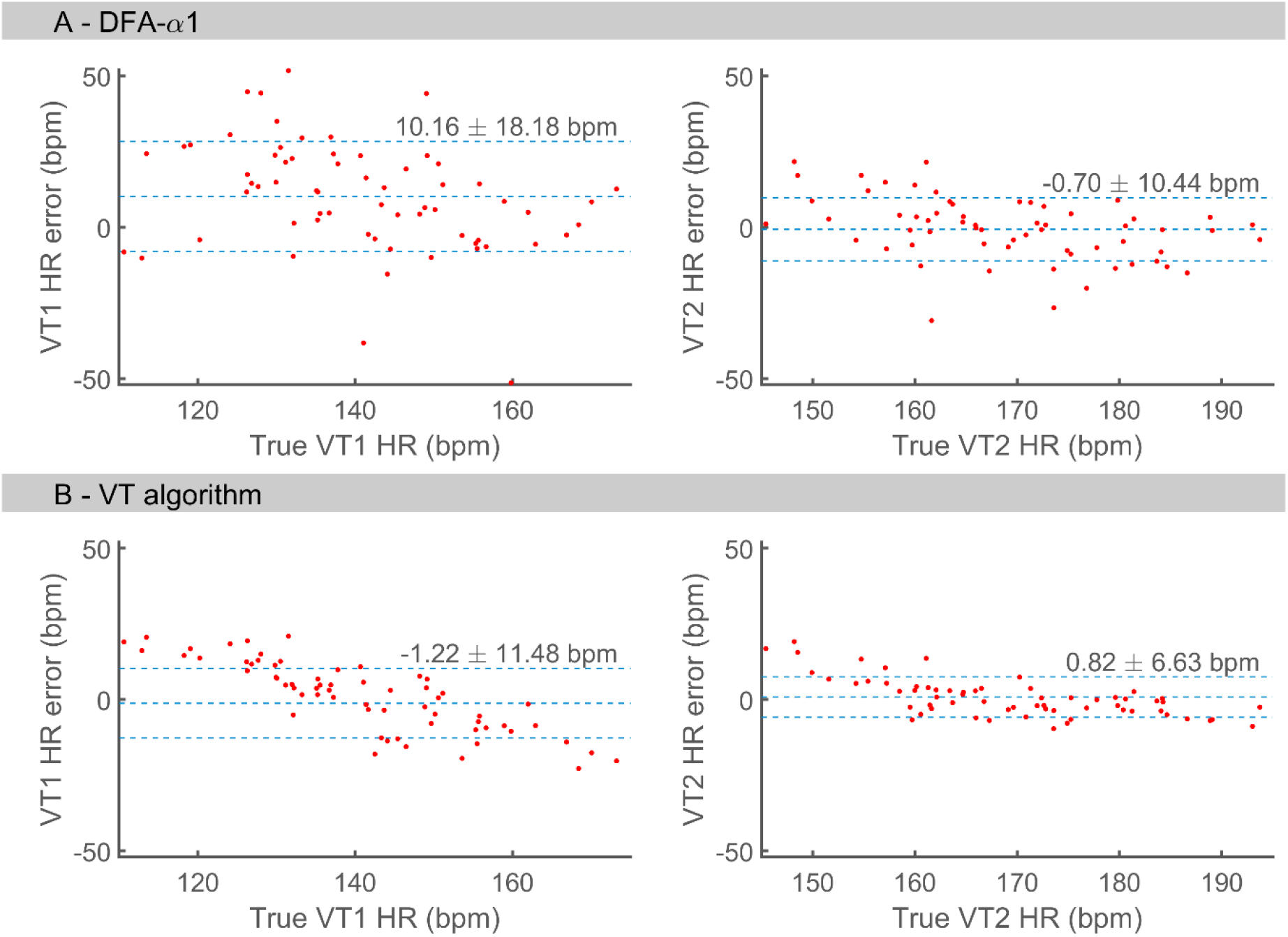
Bland-Altman analysis of HR bias and standard deviation of error (± SD) at the thresholds estimated through DFA-α1 (A) and VT-algorithm (B) compared to the true thresholds.

**Figure 3:**
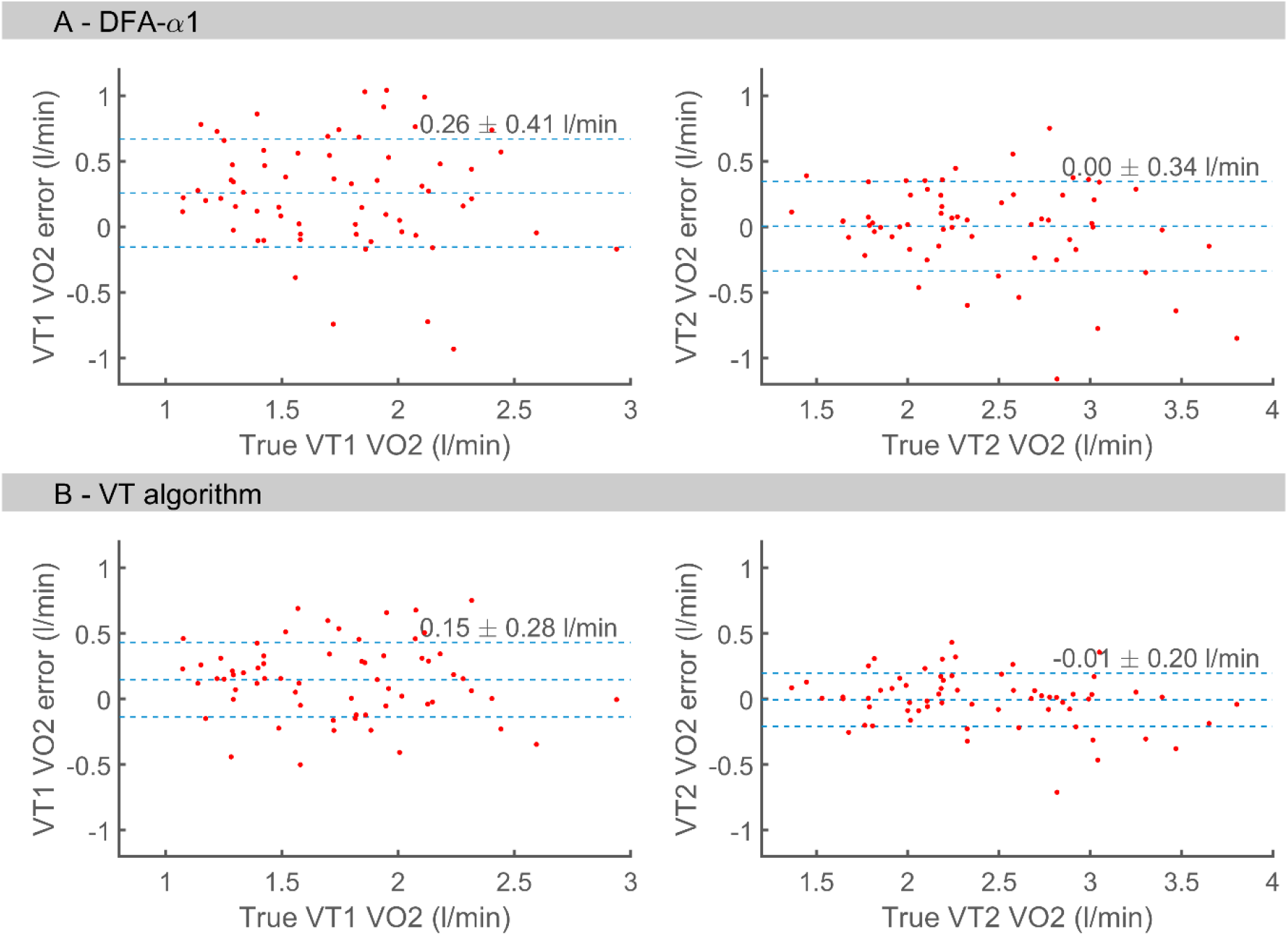
Bland-Altman analysis of VO2 bias and standard deviation of error (± SD) at the thresholds estimated through DFA-α1 (A) and VT-algorithm (B) compared to the true thresholds.

Regarding VO2 at VT1, DFA-α1 exhibited a high correlation (r=0.67, SEE 0.31) and VT-algorithm a very high correlation (r=0.81, SEE 0.26), while the mean VO2 values for both DFA-α1 (2.00 vs. 1.74 l/min, p<0.001, d=0.80) and VT-algorithm (1.89 vs. 1.74 l/min, p<0.01, d=0.40) were significantly different to the VO2 value at the true threshold. For VO2 at VT2, DFA-α1 demonstrated a very high (r=0.82, SEE 0.26) and VT-algorithm almost perfect (r=0.93, SEE 0.21) correlation, with no significant differences in the mean VO2 values determined by DFA-α1 (2.41 vs. 2.40 l/min, p=0.57, d=0.07) and VT-algorithm (2.40 vs. 2.40 l/min, p=0.59, d=0.07) when compared to the true value. Bland-Altman error statistics regarding the VO2 values at the thresholds for DFA-α1 were 0.26±41 l/min at VT1 and 0.00±0.34 l/min at VT2. For VT-algorithm, the errors were 0.15±0.28 l/min at VT1 and 0.01±0.20 l/min at VT2.

In terms of the distinctions between DFA-α1 and the VT-algorithm, both threshold estimates yielded similar results at VT2 for HR (t-test, p=0.268, d=0.14) and for VO2 (p=0.209, d=0.16). However, at VT1, significant differences were observed in both HR (p<0.001, d=0.59) and VO2 (p<0.001, d=0.59) between DFA-α1 and the VT-algorithm.

## DISCUSSION

The main purpose of this study was to validate a commercially available algorithm for accurate HRV based threshold detection. The Kubios VT-algorithm utilizes instantaneous heart rate in relation to HR reserve as well as instantaneous respiratory rate, in addition to DFA-α1, in the detection of ventilatory thresholds. We observed that due to these additional physiological inputs, the VT-algorithm was able to enhance the detection of both ventilatory thresholds when compared to the DFA-α1 approach.

The DFA-α1 algorithm, introduced by Rogers and his colleagues (7,8) has demonstrated its superiority over other methods for detecting ventilatory thresholds using HRV parameters. Similarly, as DFA-α1 algorithm also the VT-algorithm only requires an HRV recording, since both HR reserve and RF are extracted from the HRV data. Rogers and his coworkers (10,29) investigated DFA-α1 algorithms in runners via treadmill to evaluate ventilatory thresholds during exercise. Prediction of ventilatory threshold HR using DFA-α1 analysis showed promise, with VT1 HR having correlation of 0.78 and bias of 4 beats/min (error SD: ±10 beats/min) while VT2 correlation 0.93 and a bias of 2 beats/min (±5 beats/min). Especially for VT1 Mateo-March et al. (15) found high levels of validity and agreement on elite cyclists lactate thresholds. In our study, we observed notably lower correlations among DFA-α1 threshold HR values compared to prior findings. Moreover, sole DFA-α1 seems to overestimate VT1 HR by 10 beats/min and accuracy was diminished as the standard deviation of error was ±18 beats/min, though VT2 HR accuracy remained consistent with previous studies (Figure 2 A) showing a minor underestimation by 0.7 beats/min (±10 beats/min). Schaffarczyk et al. (13) reported a possible effect of sex on DFA-α1 derived VT1. However, we found no statistical differences between males and females on DFA-α1 threshold detection. This suggests that with a larger and perhaps also more heterogenous dataset, sole reliance on DFA-α1 for VT1 detection may result in reduced accuracy as also demonstrated by Fleitas-Paniagua and his colleagues (28). It is also notable that exercise modality could have an influence in the differences compared to the work of Rogers and his coworkers (10,29).

By incorporating knowledge of HR reserve and RF, the VT-algorithm appears to enhance the accuracy of threshold detection for both ventilatory thresholds compared to exclusive DFA-α1 analysis (Figure 2 and 3). The role of respiratory frequency in VT detection was investigated in a recent study by Rogers and his colleagues (18). They found that combining DFA-α1 and RF measures improved the correlation for threshold HR detection and reduced bias in HR values corresponding to both ventilatory thresholds and is in line with our results.As age-related HRmax tends to underestimate true HRmax (30) we also conducted separate analyses using age-related HRmax estimates for HRR and found no statistical difference between sole DFA-α1 and the VT-algorithm in threshold detection. Furhermore, since resting HR was not available for all study participants, a semi-ideal HRR was adopted. Despite the limitation of using a fixed resting HR in the computation of HRR, the VT-algorithm appears to enhance correlations and reduce bias, achieving at least moderate agreement with the actual measured ventilatory thresholds. However, there is an observable bias where threshold HR is overestimated at lower HR values and underestimated at higher HR values, especially seen for VT1. Whether this bias would be reduced by utilizing true HRR of not, should be studied more extensively in the future. Nonetheless, the VT-algorithm notably enhances precision in VT detection, especially concerning VT1. Both methods offer non-invasive, user-friendly analysis tools for accessing ventilatory thresholds. Rogers and his colleagues (26) validated DFA-α1 on cardiac patients, indicating strong agreement with threshold HR values.

Approximation of threshold VO2 seems to be overestimated by both DFA-α1 and VT-algorithm compared to gas exchange thresholds in VT1. However, VO2 approximation on VT2 is accurate for both DFA-α1 and VT-algorithm. These results are in pair with results by Rogers and Schaffarczyk whilst they studied VO2 relative to body mass (13,26). Consequently, both methods provide an accurate surrogate to threshold VO2 to identify exercise intensity domains and thus could be used for exercise prescription and assessing intensity distribution.

Despite the advantages of HRV-based VT detection, there are no studies employing these methods in exercise interventions. Exercise interventions typically rely on HR or VO2 values obtained from various exercise tests (e.g., CPET and the 6-minute walk test) (31,32). These tests often require expertise and access to costly laboratory equipment for lactate and VO2 measurements. Knowledge of ventilatory thresholds is essential for monitoring exercise intensity distribution and training load (33). The VT-algorithm displays nearly excellent correlation and accuracy with VO2 threshold values and, therefore, could be a valuable tool for exercise testing, exercise intensity monitoring, and training load assessment. Nowadays DFA-α1 may be used in real time monitoring (7). However, currently, the VT-algorithm is available only for post-session analysis.

## CONCLUSION

The Kubios VT-algorithm, which incorporates instantaneous HR and RF alongside DFA-α1, provides an accurate method for assessing an individual’s ventilatory thresholds. Both DFA-α1 and VT-algorithm provide an accurate estimation of different exercise intensity zones in terms of VO2. Although both methods overestimate HR in VT1 and underestimate it in VT2 the errors are lesser while using VT-algorithm compared to DFA-α1. Utilizing post-session beat-to-beat RR interval data, it provides a user-friendly and non-invasive tool for ventilatory threshold estimation. The VT-algorithm can be readily implemented in both laboratory and field settings, making it valuable for exercise testing, exercise intensity monitoring, and training load assessment.

## Data Availability

Data produced in the present study are available upon reasonable request to the authors.

## FUNDING

The current study has been funded with an award supported by the European foundation for the study of diabetes (EFSD), Juvenile Diabetes Research Foundation (JDRF) and Eli Lilly and Company (Lilly) as well as by the Finnish Diabetes Research Foundation. JAL was funded by the Research Committee of the Kuopio University Hospital Catchment Area for the State Research Funding 5101137 and 507T044.

## CONFLICT OF INTEREST

Tarvainen MP and Lipponen JA are shareholders of a company (Kubios) that designs heart rate variability analysis software. Other co-authors have nothing to declare.

## Notes

### Funding Statement

This study has been funded with an award supported by the European foundation for the study of diabetes (EFSD), Juvenile Diabetes Research Foundation (JDRF) and Eli Lilly and Company (Lilly) as well as by the Finnish Diabetes Research Foundation. JAL was funded by the Research Committee of the Kuopio University Hospital Catchment Area for the State Research Funding 5101137 and 507T044.

### Author Declarations

Ethics Committee of the Northern Savo Hospital District in Kuopio, Finland gave ethical approval for this work (484/13.00.00/2017 and 409/13.02.00/2019).

